# MetaMind: A Multi-Agent Transformer-Driven Framework for Automated Network Meta-Analyses

**DOI:** 10.1101/2025.08.04.25332893

**Authors:** Achilleas Livieratos, Maria Kudela, Yuxi Zhao, All-shine Chen, Xin Luo, Junjing Lin, Di Zhang, Sai Dharmarajan, Sotirios Tsiodras, Vivek Rudrapatna, Margaret Gamalo

## Abstract

**Background:** Network meta-analysis (NMA) enables simultaneous comparison of multiple interventions by integrating direct and indirect evidence from randomized controlled trials and observational studies, but traditional workflows require extensive manual effort for study identification, data extraction, and statistical modelling, leading to slow update cycles and operational bottlenecks.

**Objective:** To develop and validate *MetaMind*, an end-to-end, transformer-driven framework that automates NMA processes—including study retrieval, structured data extraction, and meta-analysis execution—while minimizing human input.

**Methods:** *MetaMind* integrates Promptriever, a fine-tuned retrieval model, to semantically retrieve high-impact clinical trials from PubMed; a multi-agent LLM architecture--Mixture of Agents (MoA)-- pipeline to extract PICO-structured (Population, Intervention, Comparison, Outcome) endpoints; and GPT-4o–generated Python and R scripts to perform Bayesian random-effects NMA and other NMA designs within a unified workflow. Validation was conducted by comparing *MetaMind’s* outputs against manually performed NMAs in ulcerative colitis (UC) and Crohn’s disease (CD).

**Results:** Promptriever outperformed baseline SentenceTransformer with higher similarity scores (0.7403 vs. 0.7049 for UC; 0.7142 vs. 0.7049 for CD) and narrower relevance ranges. Promptriever performance achieved 82.1% recall, 91.1 % precision and an F_1_ score of 86.4 % when compared to a previously published NMA. *MetaMind* achieved 100% accuracy in PICO element extraction and produced comparative effect estimates and credible intervals closely matching manual analyses.

**Conclusions:** *MetaMind* significantly reduces time and labour—delivering a complete NMA in under one week versus several months manually—while maintaining methodological rigor and scalability across therapeutic areas, representing a major advancement in AI-driven evidence synthesis

## Introduction

NMA is a cornerstone of evidence-based medicine (EBM), providing a robust framework for comparing multiple interventions simultaneously. By integrating both direct and indirect evidence from randomized controlled trials (RCTs) and observational studies, NMA enables researchers, clinicians, and policymakers to make informed research or treatment decisions. However, conducting NMAs is a resource-intensive process that requires significant manual effort, including study identification, data extraction, statistical modelling, and evidence synthesis.^1-4^ These demands require substantial lead time to be effective and often create operational bottlenecks, which in turn limit the timely availability of up-to-date network meta-analyses across many therapeutic areas. As a result, evidence gaps persist, leading to suboptimal treatment decisions for patients and hindering advancements in precision medicine. The lack of expertise and the extensive labour required to perform NMAs have contributed to a slow update cycle for many therapeutic areas. The challenges associated with NMA workflows—including data heterogeneity (from a population and research design/operation), bias assessment, and the need for consistent data structuring—further complicate the process.^1-4^

Artificial intelligence (AI) has the potential to transform NMA workflows by automating many of the manual processes involved, from study identification to data extraction, analysis, and interpretation. Advances in large language models (LLMs) and transformer-based retrieval systems have demonstrated remarkable capabilities in natural language processing, medical literature synthesis, and structured data extraction.^5-7^ AI-driven automation could alleviate the burdens associated with traditional NMA workflows, making the process faster, more accurate, and scalable.^1-4^ However, despite these advancements, there is currently no fully integrated, end-to-end software solution for conducting NMAs across therapy areas in a streamlined, reproducible, and flexible manner.^1-4^ Most existing AI-assisted approaches focus on isolated tasks, such as data extraction or statistical modelling, without offering a holistic pipeline that seamlessly integrates all stages of the NMA process.

Here we describe and validate *MetaMind*, a new method for automating NMAs end-to-end with minimal human input. This approach integrates a transformer-based retrieval system, Promptriever, with a multi-agent large language model framework to automate study retrieval, data extraction, and meta-analysis execution. This novel approach significantly reduces the manual effort required while improving the accuracy and scalability of NMAs. We illustrate and validate its application in a comparative efficacy study for Ulcerative colitis and Crohn’s disease, demonstrating its robustness, adaptability, and clinical utility. By streamlining the entire NMA process, our methodology represents a major advancement in AI-driven evidence synthesis, paving the way for broader applications in clinical research and health technology assessment.

While prior work has shown that structured data can be extracted from published NMAs for downstream reanalysis, such approaches are limited to post hoc extraction from already completed analyses.^1^ These methods do not address the upstream and more labor-intensive steps of the NMA lifecycle—namely, primary RCT retrieval. In contrast, *MetaMind* introduces a fully integrated, end-to-end framework built entirely in Python, which combines retrieval (Promptriever), layered MoA extraction, and dynamic Bayesian NMA script generation and execution via GPT-4o. This is the first framework, to our knowledge, to unify the entire NMA pipeline from efvidence identification through final analysis using MoA in a reproducible, extensible manner—representing a substantial methodological advancement beyond isolated automation components. *MetaMind* automates the core computational stages of network meta-analysis—study retrieval, data extraction, and statistical model execution—within a unified pipeline. While not fully autonomous in areas such as feasibility assessment or model validation, it provides an extensible framework for end-to-end automation of the technical workflow, substantially reducing manual workload and turnaround time.

## Methods

This study reports the development and validation of a new method, *MetaMind*, for automating network meta-analyses of clinical studies. To validate *MetaMind*, we used it to estimate the comparative efficacy of therapies in Ulcerative colitis and Crohn’s disease and compared our results with the results of manually performed NMAs in these therapeutic areas.

### Methodological Components and Implementation

The overview of our method called *MetaMind* is described in Figure 1, and involved chaining together several modules including information retrieval, data curation, and comparative effect estimation. This study aimed to develop and evaluated an AI-driven framework for structured clinical evidence synthesis, applying advanced retrieval and extraction methodologies to comparative efficacy analysis in moderate-to-severe inflammatory bowel disease (IBD). Specifically, the approach was designed to extract and analyze clinical trial data for UC and CD using promptable PICO metrics to ensure targeted retrieval of relevant studies. To achieve this, Promptriever was employed to search and retrieve high-impact clinical trials from PubMed, focusing on studies assessing biologic and small-molecule therapies compared to placebo or active comparators. In this work, we pre-selected the relevant studies for downstream analysis for ease of comparability and validation with the manual Bayesian Evidence Generation. The MoA framework was then applied to extract key clinical endpoints, including baseline and final remission rates, sample sizes, and confidence intervals. This structured extraction, which was FLASK (Fine-grained Language model evaluation based on Alignment Skill Sets) evaluated, enabled a comprehensive evaluation of treatment efficacy across heterogeneous study designs, facilitating a scalable and generalizable methodology for automated evidence synthesis in IBD research.

**Figure 1:**
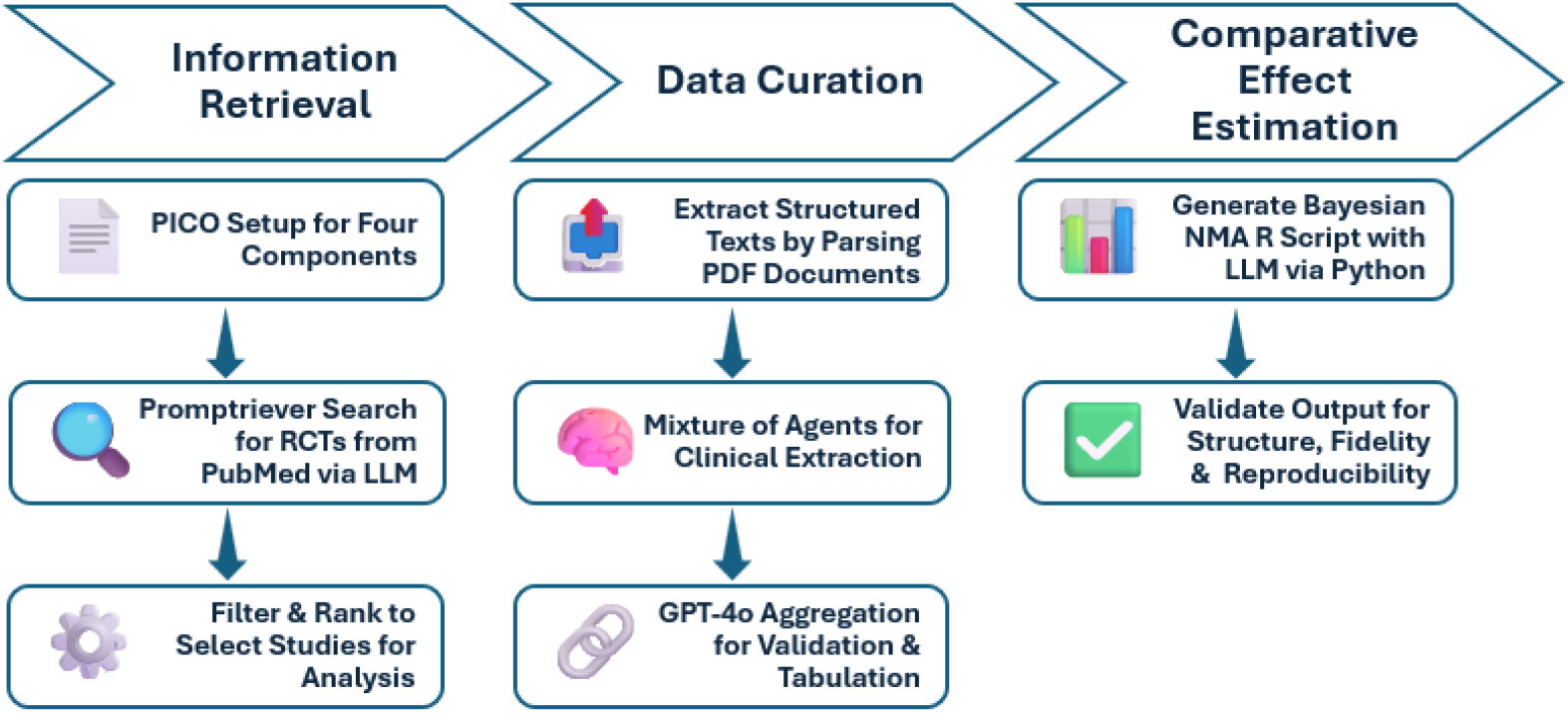
Workflow Schematic.

To operationalize PICO-aligned retrieval and extraction, we used structured, model-ready prompts tailored to each stage of the pipeline:

Retrieval via Promptriever was guided by user-defined PICO elements embedded into a query-instruction format. For example, a prompt submitted to the model might read:

> *“Query: ulcerative colitis AND placebo AND (mirikizumab OR upadacitinib OR filgotinib OR ustekinumab OR etrasimod OR tofacitinib). A relevant document would describe clinical trials where patients were tested against placebo and the document includes efficacy results for the named treatments*.*”*

This formulation enables semantic retrieval aligned with the PICO components—Population (ulcerative colitis), Intervention (named drugs), Comparator (placebo), and Outcome (efficacy).

Extraction via MoA system used standardized prompts across models like Meta-LLaMA-3, Mistral, and Qwen2 to extract structured clinical trial data. A representative example prompt is:

> *“Extract baseline and final clinical remission values for each treatment group, including confidence intervals (CIs) or standard deviations (SDs) where available. Report weekly remission rates at Weeks 4, 8, and 12. Include sample sizes per group. If any values are missing, clearly indicate and suggest plausible methods for estimation. Format output as a structured table for network meta-analysis*.*”*

These prompts enabled reliable, cross-validated extraction of quantitative endpoints from free-text PDF trial reports. More detailed examples are provided in Supplementary Figures 1 & 2.

The NMA generated through this approach was fully scripted and implemented using AI-driven automation, with GPT-4o producing the analytical script in Python. To validate the accuracy and robustness of the AI-generated NMA, results were benchmarked against a manual Bayesian Evidence Generation, conducted using R-based statistical methodologies widely accepted in evidence synthesis. This dual-implementation strategy allowed for a direct comparison between AI-assisted and traditional statistical approaches, ensuring methodological rigor and alignment with best practices in clinical research and decision-making (Figure 1).

*Metamind* workflow was then compared to a published manual NMA, to further confirm findings between the two approaches across all 3 stages of information retrieval, data curation, and comparative effect estimation.^8^

### Retrieving NMA-relevant articles using Promptriever

Promptriever is implemented in PyTorch atop a Llama-2 base Transformer with a Parameter-Efficient Fine-Tuning (PEFT) adapter. We then fixed both model and tokenizer to a maximum sequence length of 512 tokens (with padding to multiples of eight to maximize batch efficiency).

At inference time, user prompts are grouped into batches of up to four which tokenizes them with truncation and pad-to-max-length. We extract the first token embedding for each sequence—yielding a (B, 256) tensor—apply ℓ_2_-normalization across the feature dimension, and concatenate results into an (N, 256) NumPy array for all inputs.

These normalized embeddings are then compared against our library of template embeddings stored in a FAISS index. We compute inner-product similarity scores between each query vector and every template, sort in descending order, and retain the top candidates (whereby the retrieval parameter default is 100). To capture fine-grained contextual fit, each retrieved template is concatenated with the original prompt and fed to a lightweight 4-layer MLP re-ranker (512 units, ReLU activations, dropout 0.1). This network outputs a scalar relevance score for each pair.

We benchmark against the pre-trained all-mpnet-base-v2 SentenceTransformer (224 M parameters). This model generates 768-dim embeddings for both user prompts and templates. We follow the identical retrieval pipeline: compute cosine similarities over all template embeddings, sort and retain the top results (default: 100), and re-rank via the same 4-layer MLP described above. All other are kept constant to ensure direct comparability.

To facilitate a controlled validation against previously published manual NMAs, we limited the retrieval scope to a maximum of 10 high-impact articles per disease area, specifically from *The Lancet* and *NEJM*. This selection allowed for benchmarking model performance against high confidence, widely cited RCTs with standardized reporting formats. However, this constraint reflects a design choice for comparability—not a limitation of *MetaMind* itself. As shown in Supplementary, when applied without restrictions, Promptriever unrestricted application was performed against a comprehensive manual review. This demonstrates *MetaMind’s* capacity to scale beyond artificial benchmarks, enabling broader and more current real-world literature synthesis. We examined recall, precision, and F1 to evaluate our findings.

Our implementation reuses and adapts components from the Weller et al. publication, with modifications for domain-specific retrieval (Supplementary Figure 1) and integration into the NMA pipeline.^6^

### Text Extraction and Preprocessing

The next step of this methodology focused on extracting text from PDF documents, leveraging the Fitz library (PyMuPDF) to systematically gather all available textual data, such as abstracts and clinical study details. While many PubMed articles are available in machine-readable formats (XML/JSON), full-text access is often restricted due to institutional barriers, publisher policies, or non-standardized formats. PDFs present unique challenges, including inconsistent structure, embedded images, and encoding issues, making them a rigorous test case for our Mixture of Agents approach. Tables formatted as text were successfully processed; however, images, scanned tables, and graphical elements (e.g., plots) were not used in this version of the pipeline. Thus, results rely on data being presented in machine-readable textual formats. Future extensions may incorporate optical character recognition-based extraction for image-based tables or scanned documents.

By prioritizing PDF extractions, this methodology demonstrated the robustness and adaptability of our system in handling real-world document retrieval scenarios. A structured list was created, where each entry corresponded to a single PDF document, and exception handling mechanisms were implemented to address issues such as unreadable files or unsupported formats. This step was critical in preparing raw text data for subsequent semantic processing by LLMs, ensuring high-quality information retrieval even from complex, unstructured sources.

### Prompt Engineering and Layered Aggregation

The methodology employed a MoA approach, utilizing multiple LLMs across distinct inference layers to optimize clinical parameter extraction from both structured PubMed data and unstructured full-text PDFs. In the first inference layer, models including Meta-Llama-3.1-8B-Instruct-Turbo, Mistral-7B-Instruct-v0.3, and Qwen2-72B-Instruct were tasked with extracting baseline and final clinical values, standard deviations, confidence intervals, and sample sizes. To improve accuracy and consistency, the second and third inference layers employed GPT-4o, which aggregated model outputs, cross-validated extracted clinical parameters, recalculated missing values when necessary, and structured the results into a standardized format suitable for NMA. This layered approach enhanced robustness, particularly in complex PDF extractions, where inconsistent formatting, missing values, and embedded numerical data posed significant challenges. The methodology applied a recursive approach where the GPT-4o model in each layer refined the aggregated outputs from the previous iteration (Supplementary Figure 2).

### Final Synthesis

In the final stage, the responses from the aggregation layers were synthesized into a comprehensive output using GPT-4o. In this final stage, the pipeline used GPT-4o’s streaming mode to generate outputs progressively from structured prompts. All prompts were fixed in advance and applied uniformly across documents. This streaming functionality refers solely to the LLM response interface and does not imply iterative fine-tuning, editing, or adaptive prompting. Extracted data were compared post hoc to manually curated NMA reference sets for evaluation.

### PICO

The output of this methodology included a manual evaluation of PICO elements, assessing the accuracy (faithful reproduction) and completeness of the generated summaries in relation to the reference text.^2^ Rather than relying on similarity scores, which proved insufficient due to the highly detailed and comprehensive nature of the model outputs, each PICO component was individually and manually reviewed to ensure factual correctness, alignment with the reference, and clinical relevance. A qualitative assessment approach was applied, ensuring that evaluations captured nuanced differences in medical evidence synthesis.

This manual approach is particularly valuable in clinical research and systematic reviews, where accurate representation of PICO elements is essential for high-quality evidence synthesis. By prioritizing expert-driven assessment over purely computational similarity measures, the methodology ensures a rigorous and context-aware evaluation of LLM-generated summaries, setting a higher standard for automated text summarization systems in medical applications.

### FLASK

This methodology employed a multi-dimensional evaluation framework using the FLASK criteria, which assessed textual outputs based on correctness, factuality, efficiency, commonsense, comprehension, insightfulness, completeness, metacognition, readability, conciseness, and harmlessness. The evaluation process was fully automated, leveraging OpenAI’s GPT-4o to systematically score and analyse responses.^5,9^ First, the reference text was extracted from clinical study PDFs, ensuring a structured comparison between the original document and the LLM-generated response. GPT-4o was then prompted with a predefined evaluation rubric, requesting numerical scores (1 to 5) for each FLASK criterion, along with a brief justification for each score. By integrating automated scoring and structured evaluation, this framework ensured objective, reproducible, and high-fidelity assessments of LLM-generated clinical summaries (Supplementary Figure 3).

In addition to automated evaluation, we performed manual spot checks on a random sample of outputs to ensure alignment between LLM-assigned FLASK scores and human clinical interpretation. This hybrid approach helped verify that model judgments were consistent with domain-specific expectations. While LLM-based scoring enables scalable evaluation, we acknowledge that such models may carry biases from their pretraining data. To mitigate this, we used fixed rubrics and instructed the model to justify each score. This approach, along with manual spot validation, was designed to reduce variability and surface potential inconsistencies in scoring across different documents.

### Automation of NMA Generation

We used GPT-4o to write analytical code in R, using a Python-integrated workflow, to analyze the assembled dataset using brms, a validated and widely used software package for estimating comparative effects using Bayesian statistics (Supplementary Figure 4). This study employed an API-driven approach to dynamically generate and execute the code for Bayesian meta-analysis using GPT-4o.^1^ Structured pseudocode instructions were provided to GPT-4o to generate the script that defines and analyses a Bayesian random-effects model. The model evaluated relative treatment effects with a binomial likelihood and logit link function, incorporating random treatment effects at the study level and specified priors. This script was executed within Python using the subprocess module, allowing seamless integration with the remaining Python-based workflow.

More specifically, the NMA code itself was written in R by GPT-4o using structured pseudocode instructions. While the broader workflow is implemented in Python, the Python script dynamically generates and executes the R-based Bayesian model via the subprocess module. This hybrid setup was chosen to preserve the statistical rigor of established R-based packages (e.g., brms), while enabling LLM-assisted automation and integration using Python.

The generated script includes model definitions, priors, convergence diagnostics, and summary tables, and is available under Supplementary Materials.

### Bayesian Evidence Generation

The Bayesian analysis workflow was implemented using a Python-integrated workflow to evaluate treatment effects through a binomial-logit regression framework. A Bayesian random-effects model was defined with parameters for treatment-specific effects and between-study heterogeneity, using appropriately specified priors. The log-odds were estimated by modelling response counts over total sample size, incorporating treatment as a fixed effect and random treatment effects at the study level. The model output included posterior summaries with credible intervals, Rhat values for convergence diagnostics, and an estimate of between-study heterogeneity (tau). The entire analysis was executed in R-integrated Python workflow via the subprocess module, enabling seamless automation and reproducibility of Bayesian meta-analysis (Supplementary Figure 4).

In our analysis we utilized an approach within the Bayesian framework extension, which offers certain advantages like user-friendliness and familiarity over the more common NMA approach that examines treatment contrasts.^10-12^ The treatment contrast method is extensively employed in health technology evaluations, such as those conducted by the UK’s National Institute for Health and Care Excellence (NICE). The NICE decision support unit has provided methodological guidelines on the practical application of this method.^13^

### Manual R-based NMA Validation

To validate the results the manual implementation was performed for the set of selected publications.^14-23^ Standard process was followed starting with comprehensive literature search and systematic review of selected studies. To enable fair comparison between manual and automated selection, we focused on selected studies. We extracted the data on study design, interventions, and outcomes. The results derived from our manual network meta-analysis closely mirrored those obtained via our automated pipeline approach, indicating a high degree of concordance between two approaches and affirming the robustness and reliability of the automated procedure Supplementary Figure 5). In future endeavours, we plan to incorporate elements such as model evaluation, assessment of goodness-of-fit for the selected publications, and verification of underlying assumptions in the end-to-end pipeline.^24-25^

### Workflow

This entire methodology was implemented in Python, with R executed within this ecosystem, eliminating the need to rely exclusively on R or other statistical programming languages traditionally associated with NMA.^1,7^ This approach retained the strengths of R for Bayesian modelling while leveraging Python’s integration capabilities for automation, machine learning, and natural language processing. To ensure *MetaMind* meets both rigor and speed requirements, we benchmarked its entire workflow runtime against typical manual NMA workflows

### Framework for Addressing Study Heterogeneity

*MetaMind* addresses study heterogeneity through two key mechanisms: (1) the MoA framework extracts detailed trial-level data—including baseline characteristics (e.g., disease severity, prior treatment exposure, sample size, age distributions)—into a structured tabular format, and (2) the final NMA stage models between-study heterogeneity explicitly using a random-effects Bayesian model, which accounts for variance across trials. While the current pipeline does not yet implement automated subgroup adjustment or meta-regression, the structured data output allows users to identify population differences and refine inclusion or stratification rules as needed. This modular design enhances interpretability and consistency across heterogeneous study sources.

## Results

### *MetaMind* Promptriever Performance and Output

We first examined, *MetaMind* performance in retrieving relevant articles. The performance of PEFT Promptriever and SentenceTransformer was evaluated based on their similarity scores for both UC and CD. The highest similarity score achieved by PEFT Promptriever was 0.7403, marginally outperforming SentenceTransformer’s 0.7049 (Table 1a). Additionally, PEFT Promptriever demonstrated a narrower similarity range (0.0814) compared to SentenceTransformer (0.0959), indicating a tighter focus on relevant articles and higher specificity in aligning to nuanced query instructions. Similarly, on CD PEFT Promptretriever demonstrates more consistent performance with a narrower range (0.0353), focusing on high-relevance results (Table 1b). A filter was applied to select only the top 10 most relevant articles.

**Table 1a:** Performance Comparison Between Retrieval Models for Ulcerative Colitis.

| Model | Highest Similarity | Lowest Similarity | Range |
| --- | --- | --- | --- |
| PEFT Promptriever | 0.7403 | 0.6589 | 0.0814 |
| SentenceTransformer | 0.7049 | 0.6090 | 0.0959 |

**Table 1b:**
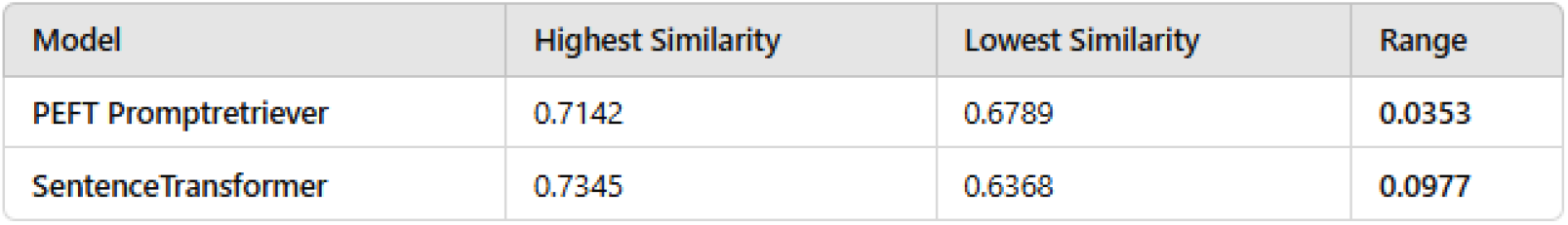
Performance Comparison Between Retrieval Models for Crohn’s Disease.

The top five articles on the induction and maintenance therapy for ulcerative colitis were filtered (Table 2a). The study on Etrasimod achieved a similarity score of 0.7043. Upadacitinib scored 0.6998 and explored advanced therapies for moderate to severe ulcerative colitis These findings highlight the capability of PEFT Promptriever to retrieve high-quality, clinically relevant studies with nuanced semantic alignment.

**Table 2a:**
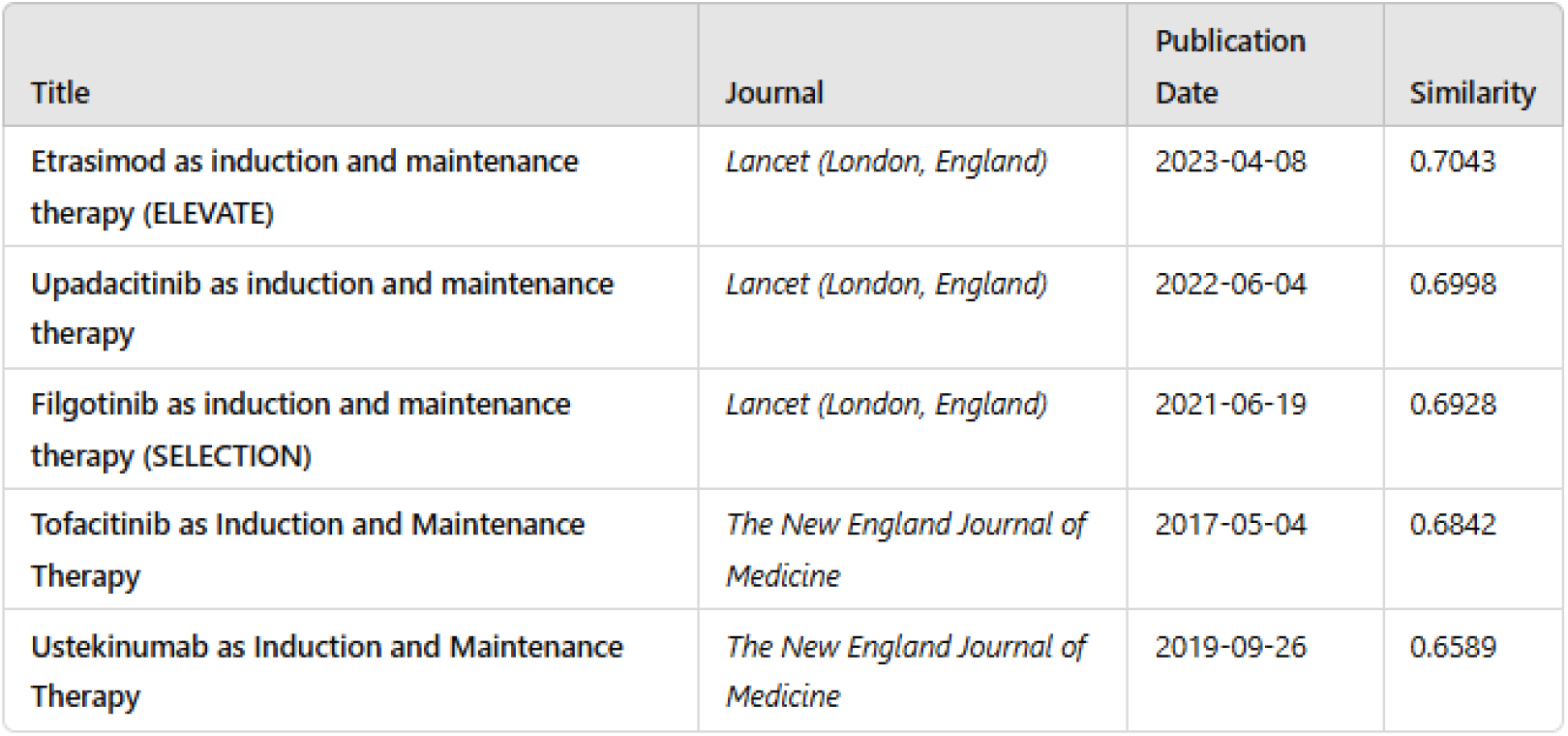
Promptriever Output-Key Ulcerative Colitis Articles Examples.

**Table 2b:**
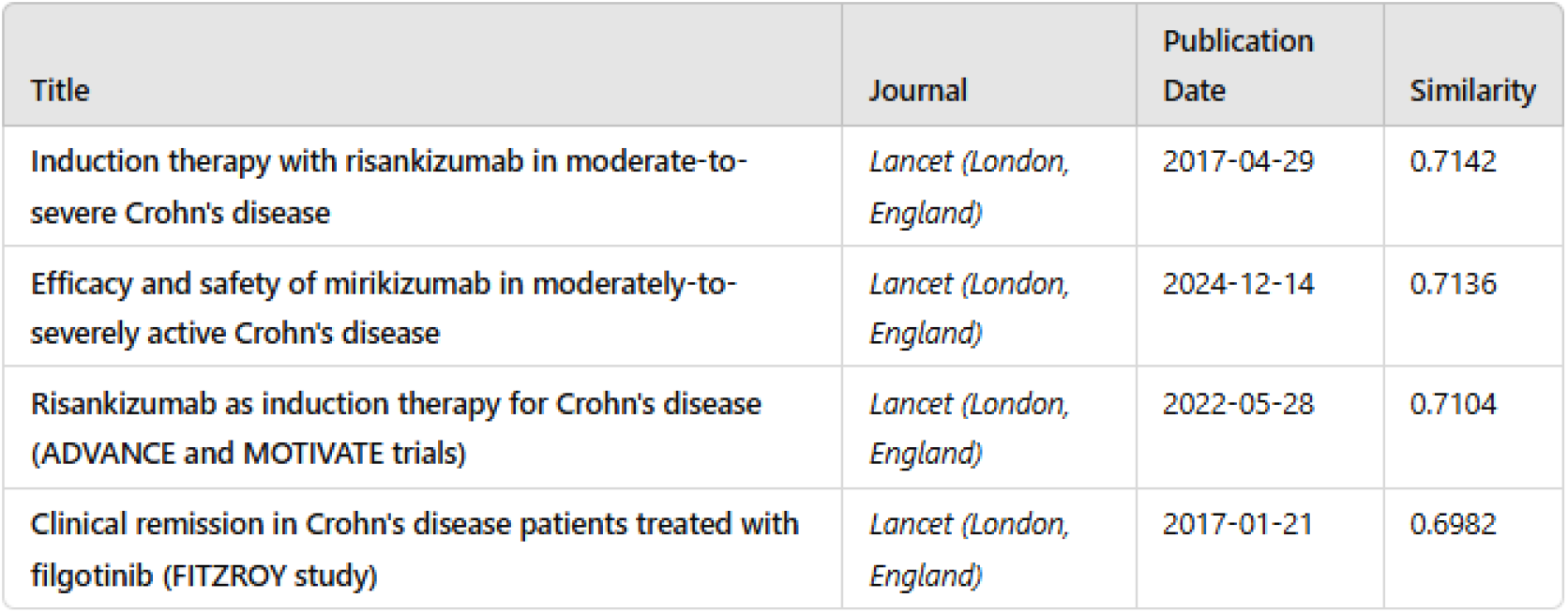
Promptriever Output-Key Crohn’s Disease Articles Examples.

| Title | Journal | Publication Date | Similarity |
| --- | --- | --- | --- |
| Induction therapy with risankizumab in moderate-to-severe Crohn's disease | *Lancet (London, England)* | 2017-04-29 | 0.7142 |
| Efficacy and safety of mirikizumab in moderately-to-severely active Crohn's disease | *Lancet (London, England)* | 2024-12-14 | 0.7136 |
| Risankizumab as induction therapy for Crohn's disease (ADVANCE and MOTIVATE trials) | *Lancet (London, England)* | 2022-05-28 | 0.7104 |
| Clinical remission in Crohn's disease patients treated with filgotinib (FITZROY study) | *Lancet (London, England)* | 2017-01-21 | 0.6982 |

A similar approach was taken for Crohn’s disease. The top four articles on the induction and maintenance therapy for Crohn’s disease were filtered (Table 2b). The study on Risankizumab achieved the highest similarity score of 0.7142 and focused on clinical remission in moderate-to-severe Crohn’s disease.

To validate Promptriever’s retrieval accuracy, we compared its results against a published NMA which included 28 relevant trials (Supplementary Figure 6). Of these, Promptriever successfully retrieved 23, resulting in a recall of 82.1%. All retrieved trials were relevant, yielding a precision of 91.1% and an F1 score of 86.4%. The 5 studies missed by Promptriever included 1 without a PubMed ID, which was not retrievable. These findings demonstrate that Promptriever delivers high-recall, high-precision semantic retrieval while also surfacing more recent and diverse studies than the manual reference set. Moreover, as not every alternative Promtriever frameworks was explored, it is plausible that due to workflow flexibility these summary statistics can be further improved.

### *MetaMind* MoA Output

*MetaMind’s* MoA module was evaluated for its ability to extract structured clinical data from unstructured PDF trial reports. For selected trials in UC and CD, the system correctly extracted numerical endpoints (e.g., remission rates, confidence intervals, and sample sizes) across two timepoints per study, demonstrating successful mapping to NMA-ready tabular format (Tables 3a & 3b). These outputs were validated against manually curated references to assess extraction fidelity.

**Table 3a:** Data Extraction for UC of Ustekinumab using MoA.

| Treatment | Time Point | Remission Rate (%) | SE | 95% CI | Sample Size |
| --- | --- | --- | --- | --- | --- |
| Ustekinumab (130 mg) | Week 8 | 15.6 | 0.0207 | (11.5, 19.7) | 321 |
| Ustekinumab (6 mg/kg) | Week 8 | 15.5 | 0.0206 | (11.4, 19.6) | 320 |
| Placebo | Week 8 | 5.3 | 0.0125 | (2.8, 7.8) | 319 |
| Placebo | Week 44 | 24.0 | 0.0327 | (17.6, 30.4) | 175 |
| Ustekinumab (90 mg every 12 weeks) | Week 44 | 38.4 | 0.0371 | (31.1, 45.6) | 172 |
| Ustekinumab (90 mg every 8 weeks) | Week 44 | 43.8 | 0.0373 | (36.5, 51.1) | 176 |

**Table 3b:** Data Extraction for CD of Upadacitinib using MoA.

| Trial | Treatment | Week 12 Remission % (CI) | Week 52 Remission % |
| --- | --- | --- | --- |
| U-EXCEL | Placebo | 29.1% (22.4–35.8) | N/A |
| U-EXCEL | Upadacitinib 45 mg | 49.5% (44.2–54.8) | N/A |
| U-EXCEED | Placebo | 21.1% (14.9–27.2) | N/A |
| U-EXCEED | Upadacitinib 45 mg | 38.9% (33.6–44.2) | N/A |
| U-ENDURE | Placebo | N/A | 15.1% |
| U-ENDURE | Upadacitinib 15 mg | N/A | 37.3% |
| U-ENDURE | Upadacitinib 30 mg | N/A | 47.6% |

We assessed the accuracy of extracted PICO elements by manually reviewing outputs from a subset of UC and CD trials. For this limited evaluation (focused on remission-related endpoints), the MoA framework correctly extracted and aligned all targeted data fields, achieving 100% accuracy on these samples. However, we acknowledge that broader validation across multiple endpoints and trial designs is needed before generalizing this performance metric further.

FLASK variability was observed due to study-level differences in reported endpoints and formatting (Supplementary Figure 3). However, the use of universal prompts across all studies was sufficient to capture all required clinical information, even in the presence of structural variability (Supplementary Figure 2). This demonstrates the generalizability and robustness of the prompt design, which enabled consistent extraction without the need for study-specific tailoring.

### *MetaMind* NMA Output

To validate the accuracy of *MetaMind’s* automated NMA pipeline, we conducted two complementary comparisons. First, we compared the AI-generated NMA results (Figures 2a and 2b) to manually performed analyses by the co-authors (Supplementary Figure 5).^14-23^ This internal benchmark showed high concordance in effect estimates, credible intervals, and treatment rankings, confirming the reliability of *MetaMind’s* outputs. Second, we assessed alignment with a recent publication, a peer-reviewed NMA in ulcerative colitis (Supplementary Figures 6 and 7).^8, 26-33^ This demonstrates consistent treatment effect patterns and overlap in identified trials, supporting external validity. Taken together, these findings confirm that *MetaMind* replicates both internal and published NMA outputs with high fidelity.

**Figure 2:**
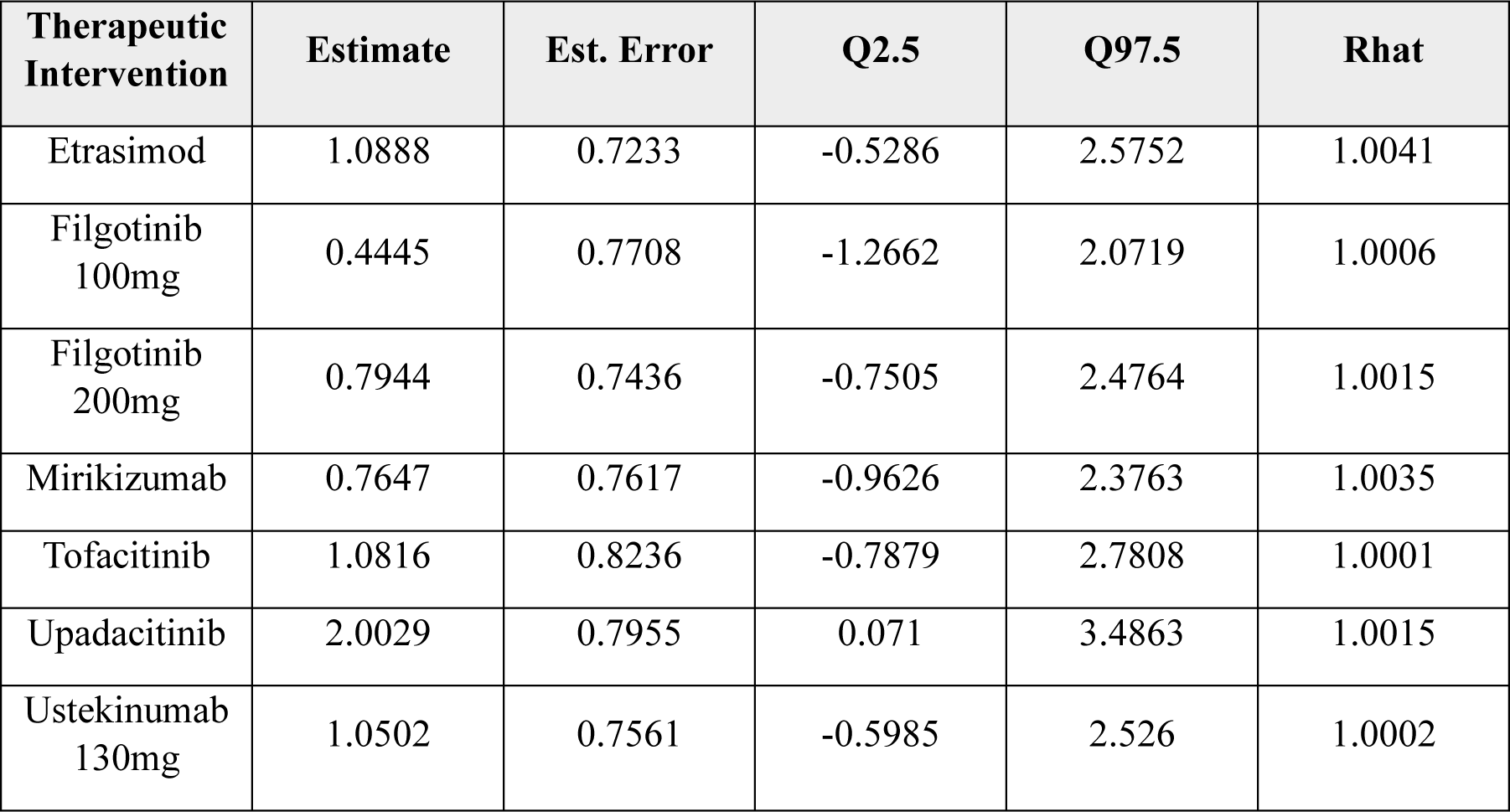
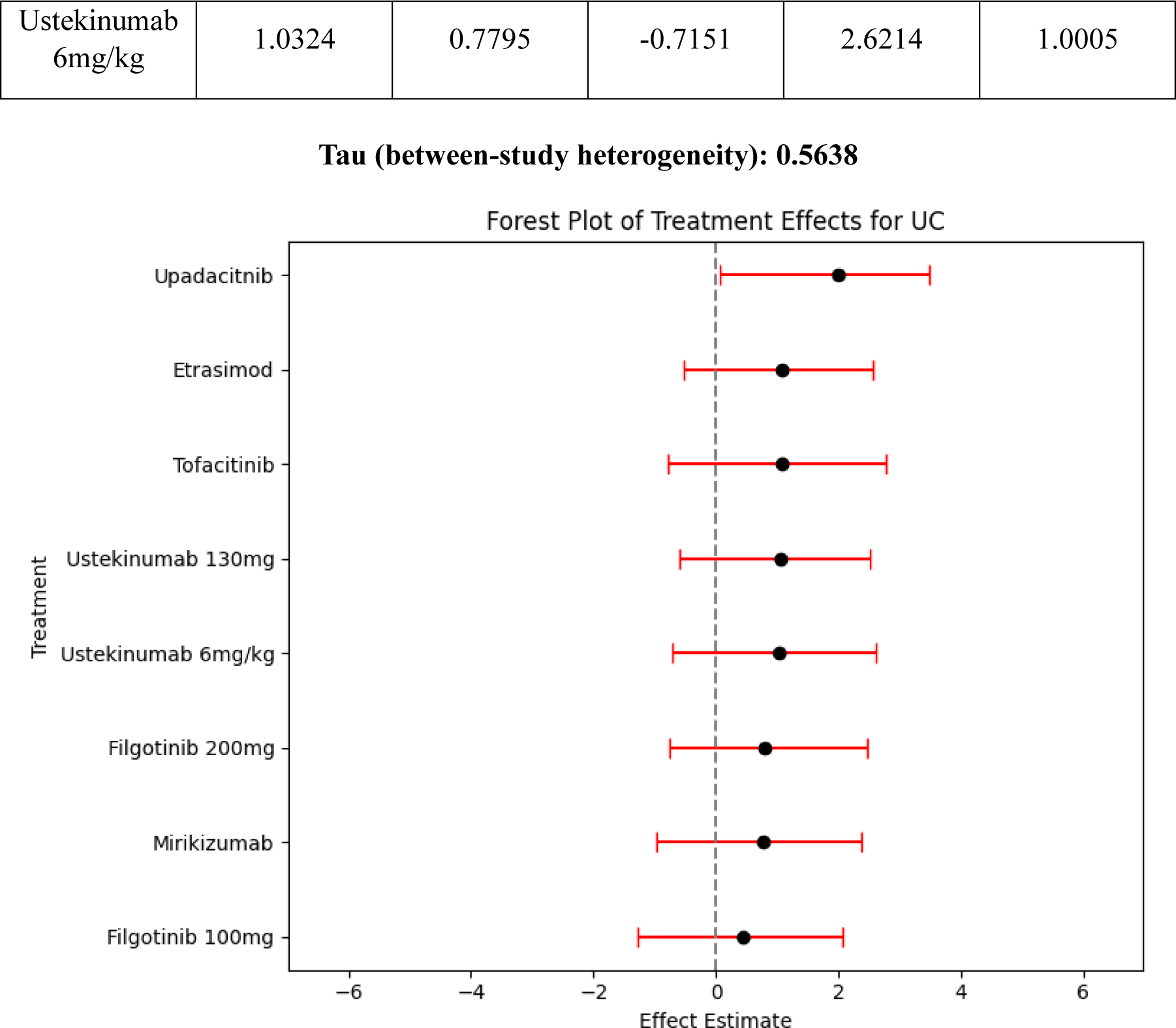
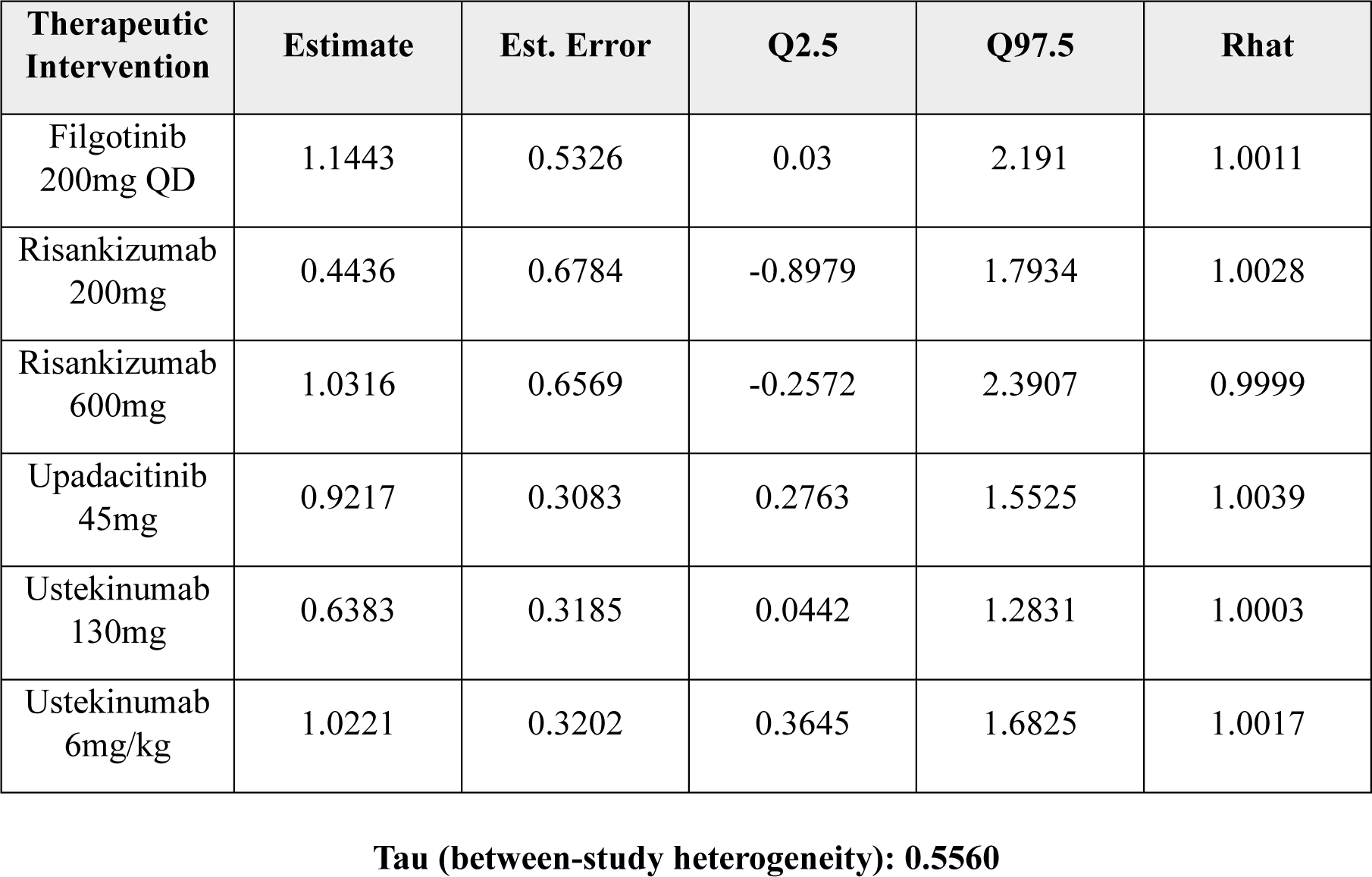
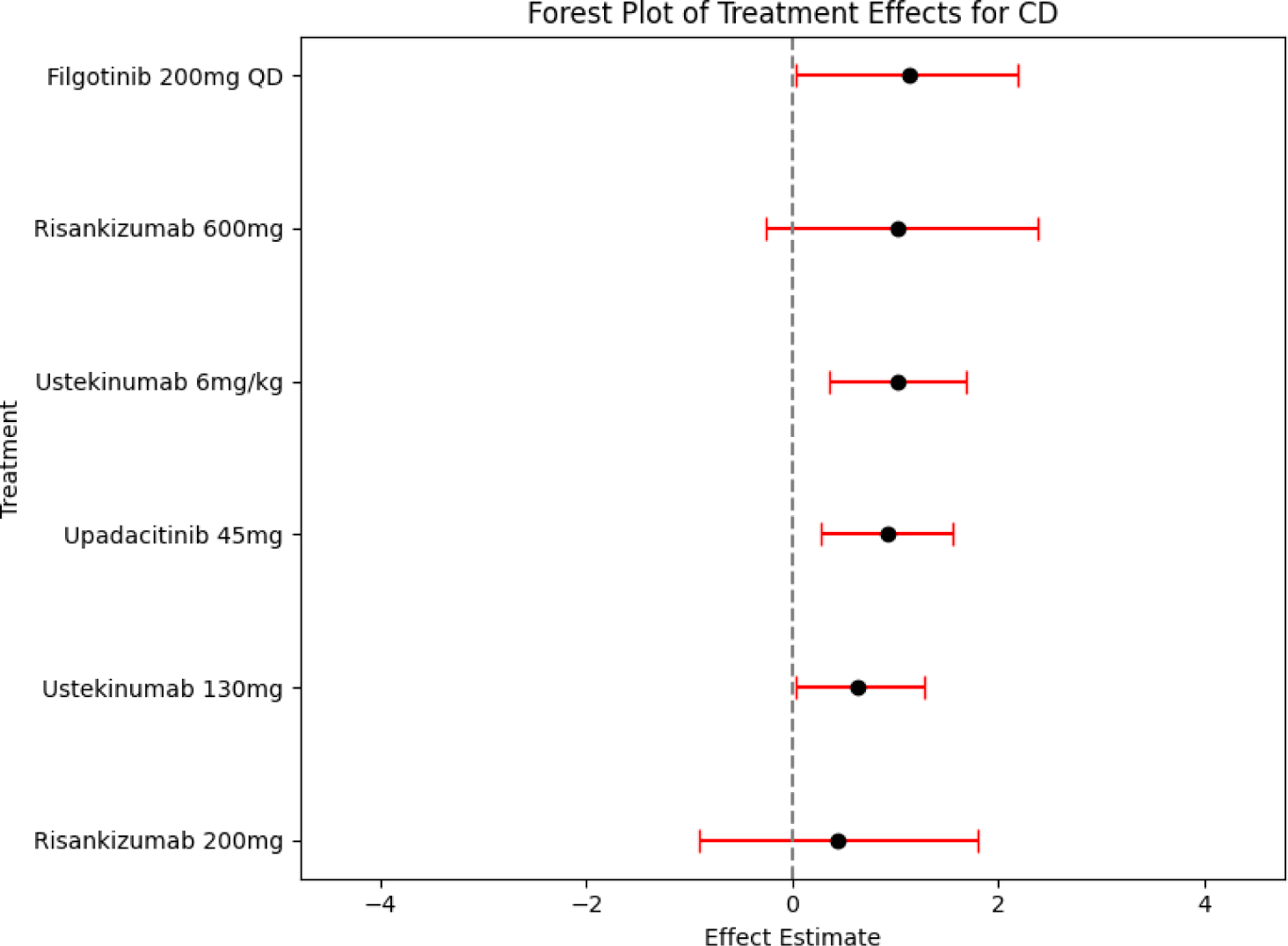
(a) UC NMA Summary & Forest Plot (b) CD NMA Summary & Forest Plot.

Finally, *MetaMind* outputs can be generated within a week, while the entire workflow to be generated manually would require several months.^34^

A stage-wise comparison of *MetaMind* against manual NMA workflows is summarized in Table 4, highlighting performance across retrieval accuracy, data extraction fidelity, statistical concordance, and execution time.

**Table 4:** Summary table of *MetaMind* performance across all stages.

| Pipeline Stage | Metric | *MetaMind* Result |
| --- | --- | --- |
| Retrieval | Recall (vs manual NMA) | 82.1% |
|  | Precision | 91.1% |
| Data Extraction | PICO element accuracy | 100% |
| NMA Concordance | Treatment overlap (UC/CD) | Comparable-No meaningful differences |
|  | Tau, Rhat, CI diagnostics | Consistent with manual implementation |
| End-to-End Time | Total workflow duration | < 1 week (automated) vs ~3–4 months |

## Discussion

The integration of advanced AI methodologies such as Promptriever and multi-agent architectures into NMA workflows signifies a shift in clinical evidence synthesis. The results underscore the efficiency of Promptriever’s PEFT-enabled retrieval system in surfacing highly relevant and nuanced PubMed studies, with evaluation metrics surpassing conventional retrieval models. By seamlessly coupling this retrieval mechanism with a multi-agent LLM framework, the pipeline ensures rigorous data extraction and synthesis, delivering structured and actionable insights with minimal human oversight. This workflow not only streamlines traditionally labour-intensive NMA processes but also enhances their scalability and adaptability across diverse therapeutic areas.

This study not only seeks to enhance the efficiency of NMA processes but also aims to establish a replicable and robust framework for integrating AI-driven methodologies in broader evidence synthesis tasks.^1−2,5^ The inclusion of open-source models within a multi-agent architecture ensures adaptability and customization, which are essential for meeting the diverse needs of healthcare research.^2^ Our prompt engineering techniques address limitation of generalizability by crafting highly specific instructions and contextual inputs for LLMs, thereby reducing the need for extensive manual oversight while improving scalability and reliability.^5,9^ Moreover, by retrieving a broader and more up-to-date set of trials than a recent peer-reviewed manual NMA (Supplementary Figure 6), *MetaMind* demonstrates its real-world applicability and capacity to exceed traditional methods in both scope and timeliness. Importantly, *MetaMind* is not a proprietary tool, but rather a reproducible research workflow built on open-source components and publicly available models. It is designed to be transparent, modifiable, and extensible for future researchers seeking to implement automated NMAs without relying on commercial software. Although *MetaMind* automates the primary data and modeling stages of NMA, it does not yet perform upstream feasibility assessment, eligibility screening, or statistical model selection based on fit or inconsistency checks. These remain essential expert-guided tasks. Thus, our use of the term “end-to-end” refers specifically to the computational execution pipeline, not to full autonomy across all stages of NMA methodology.

In conclusion, this workflow’s modularity, adaptability, and accuracy offer a leap forward in automated evidence synthesis. The results from Promptriever’s PEFT-enabled retrieval in Step 1, the layered multi-agent extraction in Step 2, and the automated code generation and NMA execution in Step 3 exemplify a system designed to scale with evolving datasets, therapeutic areas, and analytical demands within a unified workflow.

### Limitations

Despite its strengths, the workflow has notable limitations. First, the dependency on pre-trained LLMs such as GPT-4o raises concerns about transparency and reproducibility, particularly in closed-source environments. While incorporating open-source models mitigates vendor lock-in, it still necessitates ongoing updates and fine-tuning to maintain relevance across shifting clinical domains. Additionally, while Promptriever’s PEFT adaptation optimizes computational efficiency, it may still face challenges when addressing highly heterogenous datasets or rare conditions, which require more extensive contextual understanding. The retrieval process was designed to prioritize highly relevant studies, but in doing so, some potentially useful papers may have been excluded. The search parameters were structured to maximize precision over recall, ensuring that the selected studies were of high relevance. Future iterations could employ broader, multi-stage searches, ensuring a more comprehensive dataset while maintaining retrieval precision.

Moreover, the reliance on static pre-trained embeddings limits the pipeline’s real-time adaptability to newly emerging clinical evidence. This constraint is particularly evident in areas with rapidly evolving treatments, where retraining models may introduce delays. The robustness of Bayesian NMA outputs could also be impacted by the quality of initial inputs, necessitating rigorous pre-processing and curation. This work does not aim to establish or refine specific statistical models but rather to demonstrate the feasibility of an end-to-end automated pipeline. The focus is on proving that AI can successfully retrieve clinical studies, extract structured data, and execute NMAs with minimal human intervention. While the AI pipeline effectively automates these tasks, domain experts are still needed to validate statistical models, adjust assumptions, and interpret results in context. Further work should incorporate a wider search strategy and develop rule-based classifiers for eligibility screening, enabling the pipeline to both discover and autonomously investigate all potentially relevant trials.

### Future Directions

To address these limitations, future research could explore the integration of agentic LLMs— autonomous AI agents capable of dynamically adapting to complex workflows.^35^ Agentic systems would enhance scalability by iteratively refining models based on real-time feedback, offering improved contextualization and responsiveness to emerging clinical data. Additionally, integrating quantum computing into the pipeline could revolutionize computational efficiency, enabling faster data processing and retrieval for high-dimensional datasets.

Quantum computing’s potential for parallel processing aligns well with the demands of Bayesian NMA, where multidimensional treatment comparisons are computationally intensive. Quantum-based models could also advance semantic retrieval by leveraging quantum-inspired algorithms for nuanced similarity scoring, further reducing noise and enhancing precision in evidence synthesis.^36^

Finally, expanding the workflow to incorporate adaptive learning mechanisms and multilingual capabilities would make it more globally applicable, particularly in low-resource settings. By combining these advancements, the workflow could set new standards for automated clinical analytics, enabling broader accessibility and real-time adaptability in evidence synthesis tasks.

## Acknowledgment

No further acknowledgements

## Authorship Confirmation

All authors certify that they meet the ICMJE criteria for authorship.

## Funding/Support

The authors received no financial support for this research.

## Role of the Funder/Sponsor

The funder had no role in study design; data collection, analysis, or interpretation; manuscript preparation; or decision to submit for publication.

## Author Approva

All authors have seen and approved the manuscript

## Data Availability Statement

No new data were created or analysed in this study. Data sharing is not applicable to this article

## Author Information Competing Interests

A.C., M.K., X.L., M.G., and Y.Z. are employees of Pfizer Inc. D.Z. is an employee of Teva Pharmaceuticals USA. J.L. is an employee of Takeda Pharmaceuticals USA. S.D. is an employee of Sarepta Therapeutics. S.T. has received institutional research funding from the ECRAID-Base consortium funded by the EU Horizon 2020 programme. V.R. has received research funding from BeOne Medicines Ltd; during the past 36 months has received contracts or grants from Blueprint Medicines, Genentech, Janssen, Merck, Mitsubishi Tanabe, Stryker, and Takeda; received honoraria from Natera and Ironwood; and served in leadership roles with Data Unite, ZebraMD, and AcucareAI. A.L. declares no conflicts of interest.

## Author Contributions

M.G. and A.L. conceived the idea; A.L., M.K., Y.Z., A.C., J.L., D.Z., M.L., S.D., S.T., V.D. and M.G. performed the literature search; A.L., M.K., Y.Z., A.C., J.L., D.Z., M.L., X.L. and S.D. performed the analysis; A.L., M.K., Y.Z., S.T., V.D. and M.G wrote the manuscript; S.T., V.D. and M.G. critically corrected the manuscript; M.G. oversaw the study. All authors have read and agreed to the published version of the manuscript.

## Supplementary Material

**Supplementary Figure 1: Promptriever Code**

**Placeholder**

**Supplementary Figure 2: Mixture of Agents Code**

**Placeholder**

**Supplementary Figure 3:**
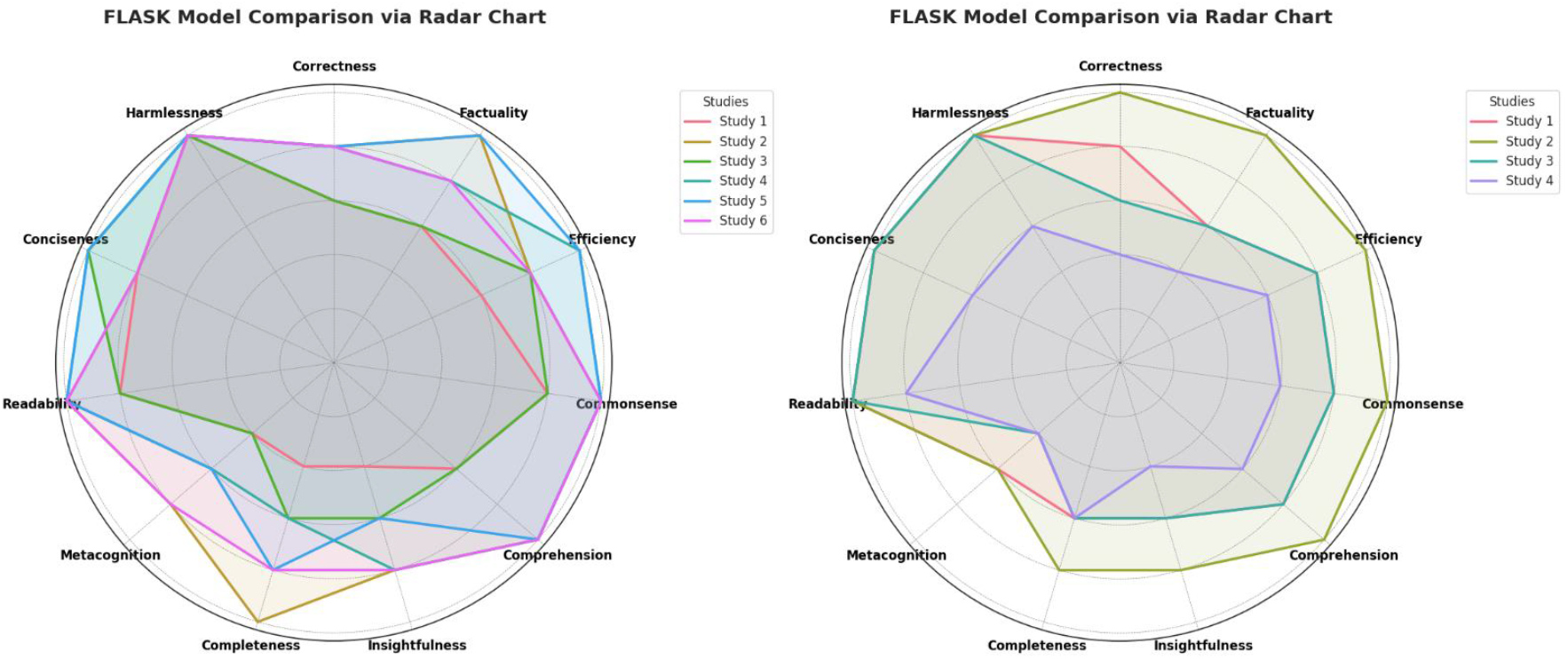
FLASK UC and CD Plots. Variability in FLASK metrics is associated with broad prompts not always applicable for each publication.

**Supplementary Figure 4: Code-NMA Script Generation**

**Placeholder**

**Supplementary Figure 5:**
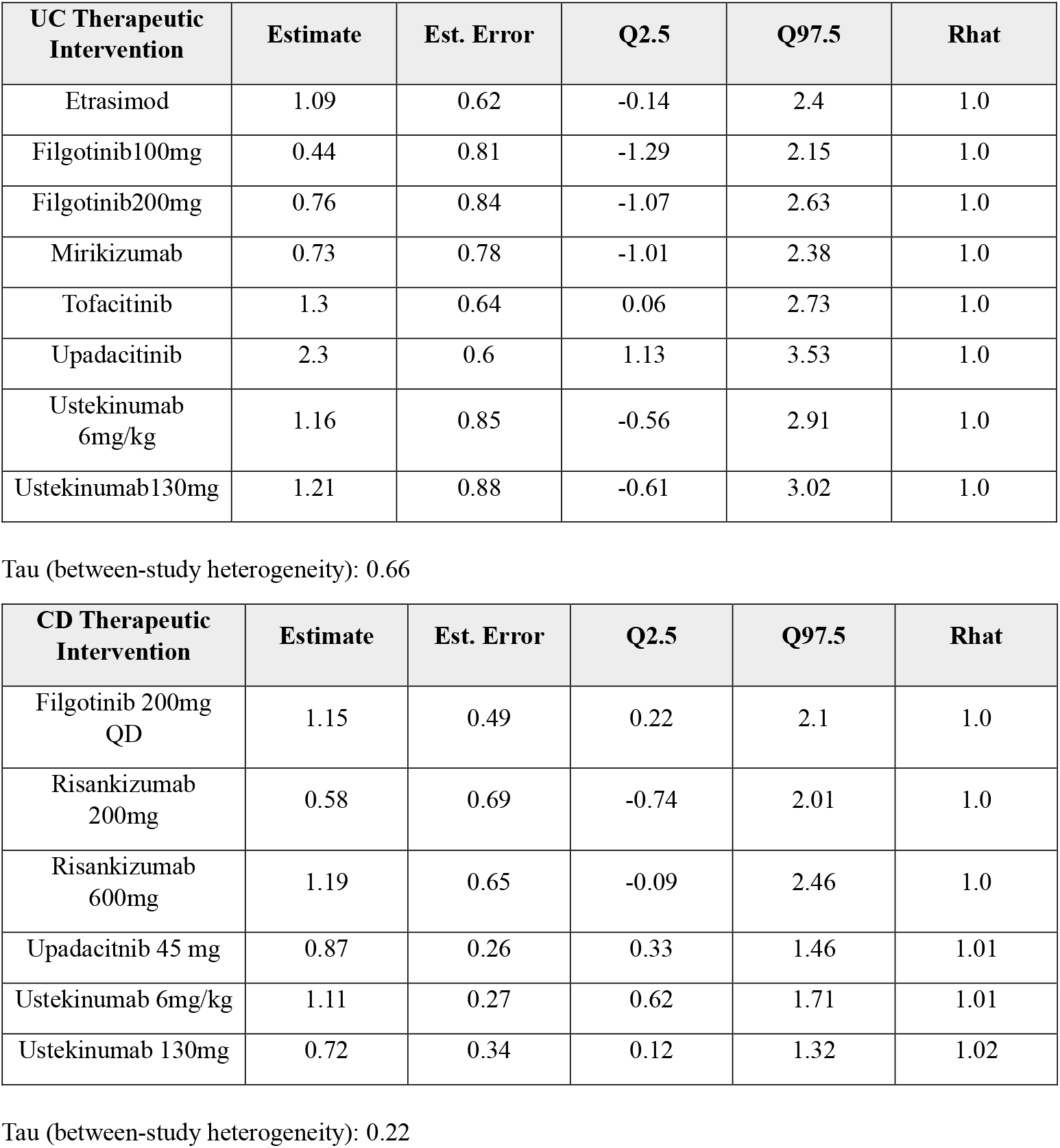
UC and CD Manual NMA Validation.

**Supplementary Figure 6:**
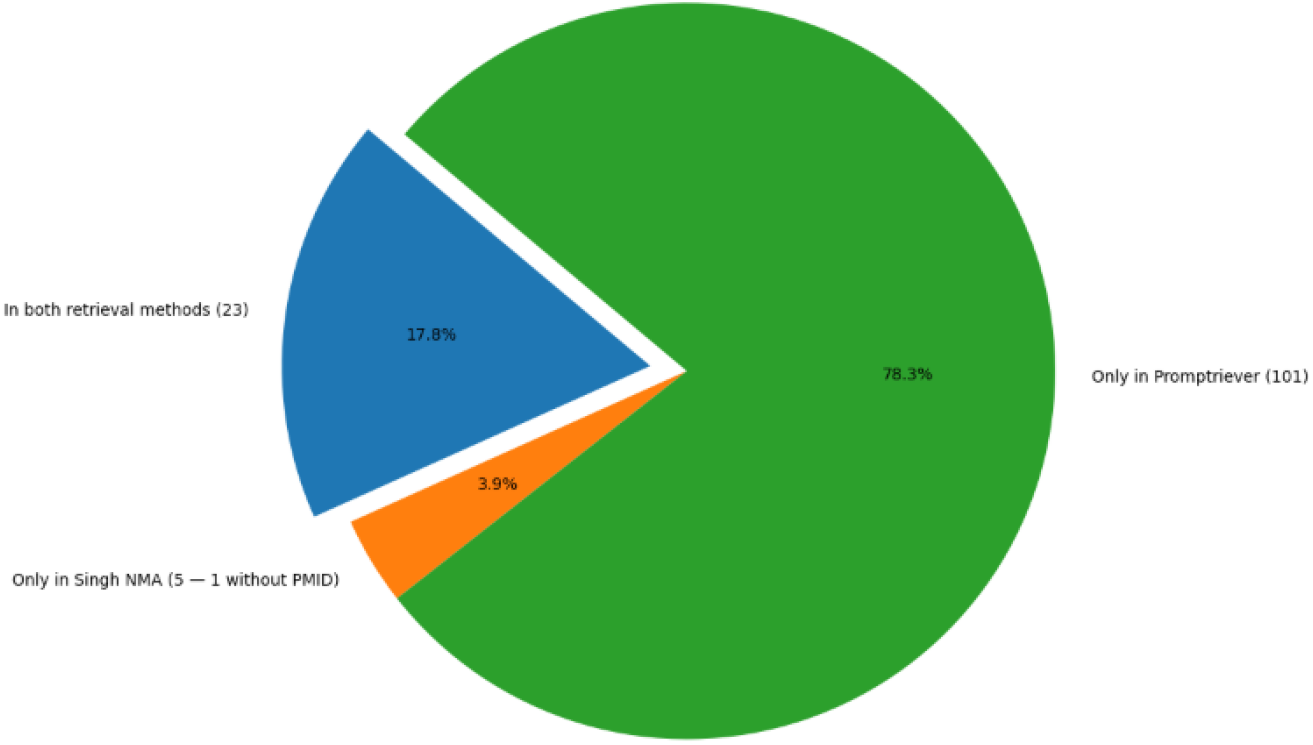
Information Retrieval using Promptriever versus Published NMA. Promptriever recovered the vast majority (78.3%) of studies not captured by Singh et al.’s manual NMA—many of which were recent PubMed-indexed trials or combination-therapy investigations that post-dated the original review. Only 3.9% of trials were uniquely in the published NMA (and 1 study not PubMed-indexed), while 17.8% appeared in both approaches. This suggests that Promptriever not only replicates almost all of the published NMA’s PubMed sourced evidence, but also efficiently surfaces the newest and most diverse treatment studies beyond the scope of the manual search.

**Supplementary Figure 7:**
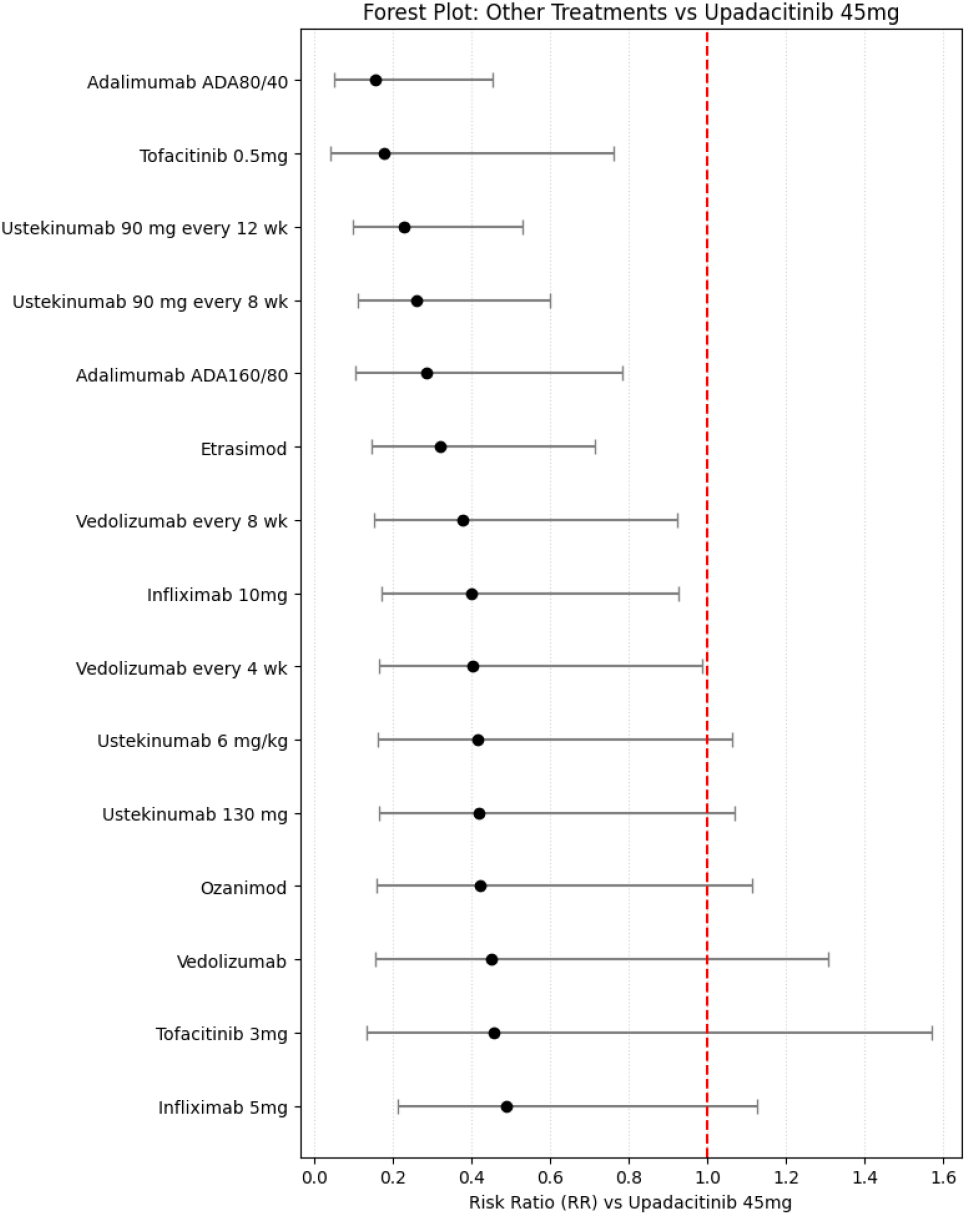
NMA Generation using *Metamind*. Note: NMA generated from a total of 8 RCT studies

**Supplementary Figure 8: Code-NMA Script Execution**

**Placeholder**

